# Variation in COVID-19 Outbreaks at U.S. State and County Levels

**DOI:** 10.1101/2020.04.23.20076943

**Authors:** Wolfgang Messner, Sarah E. Payson

## Abstract

**Background:** The COVID-19 pandemic poses an unprecedented threat to the health and economic prosperity of the world’s population. Yet, some countries or regions within a country appear to be affected in different ways.

**Objectives:** This research aims to understand whether the outbreak varies significantly between U.S. states and counties.

**Methods:** A statistical model is estimated using publicly available outbreak data in the U.S., and regional differences are statistically analyzed.

**Results:** There is significant variance in outbreak data between U.S. states and counties. At the state level, the outbreak rate follows a normal distribution with an average relative growth rate of 0.197 (doubling time 3.518 days). But there is a low degree of reliability between state-wide and county-specific data reported (ICC = 0.169, *p* < 0.001), with a bias of 0.070 (standard deviation 0.062) as shown with a Bland-Altman plot.

**Conclusions:** The results emphasize the need for policy makers to look at the pandemic from the smallest population subdivision possible, so that countermeasures can be implemented, and critical resources provided effectively. Further research is needed to understand the reasons for these regional differences.

## Introduction

On January 20, 2020, the first case of the novel coronavirus disease 2019 (COVID-19) was reported on U.S. soil. Original efforts to contain the virus were ineffective, with cases in the U.S. growing to over 579197 as of April 13, 2020.^1^ In the struggle to contain the pandemic’s growth rate, the U.S. Government has taken unprecedented action, imposing international and domestic travel restrictions, closing down businesses, and enacting state-wide stay at home orders and social distancing mandates. These steps are intended to not only slow the spread of the virus, but to minimize peak levels to prevent exhausting necessary resources and hospital capacities.

A community’s susceptibility to any virus is determined by a variety of factors, including but not limited to biological determinants, demographic profiles, and socioeconomic characteristics.^2^ Because these factors vary significantly across the U.S., there is likely to be considerable intra-country variation in the outbreak as well. In fact, the local popular press is reporting such differences, sometimes between neighboring counties.^3^ Because country and state-level epidemiological estimation models can potentially hide such local dynamics, policy makers have been advised to look at the pandemic from the smallest population subdivision possible in order to understand its progression, the effectiveness of countermeasures, and the need to provide critical medical resources, such as hospital beds, intensive care unit beds, and ventilators.^2^

In the current study, we examine the relative growth rate of the COVID-19 outbreak and its variation on a state and county level across the U.S. We show, both through visualizations as well as statistical analysis, that the outbreak varies significantly across counties and that an aggregate view at the state level, as it is most often reported in media, hides differences at a lower level. Additionally, while data are collected within local medical facilities during an emerging pandemic, outbreak rates are typically reported at aggregate levels.^4^ In this article, we show the necessity of analyses on a lower level.

This paper proceeds as follows. In the next section, we detail our data sources and the statistical estimation model. Then we present our statistical results, both at country, state, and county level. Finally, we discuss the implication of the findings and outline our further research in this area, which will be aimed at understanding the structural factors driving the variation.

## Model and methods

We obtain COVID-19 outbreak data from the China Data Lab published at Harvard Dataverse (as of April 13, 2020) and USA Facts (as of April 14, 2020),^1,5^ and check for consistency between the two databases. Since January 22, 2020, the latter database has aggregated data from the Centers for Disease Control and Prevention (CDC) and state- and local-level public health agencies, confirming them by referencing state and local agencies directly. The 21 cases confirmed on the Grand Princess cruise ship on March 05 and 06, 2020 are not attributed to any counties in California. For our county-level analysis, we discard cases which USA Facts can only allocate at the state, but not at the county level due to a lack of information. On average, the number of unallocated cases is small, but a few states contribute as many as 4866 (New Jersey), 1300 (both Rhode Island and Georgia), or 1216 (Washington State) unallocated cases, resulting in an average of 308 unallocated cases per state, again as of April 14, 2020.

Following approaches by the Institute for Health Metrics and Evaluation at the University of Washington^6^ and the COVID-19 Modeling Consortium at the University of Texas at Austin,^7^ we statistically model the outbreak in the U.S. at state and county level using the exponential growth equation 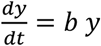, where *b* is a positive constant called the relative growth rate; it has units of inverse time. Going forward, we simply refer to *b* as the outbreak rate. Solutions to this differential equation have the form *y* = *a e*^*bt*^, where *a* is the initial value of cases *y*. The doubling time *T*_*d*_ can be calculated from the exponential growth rate as 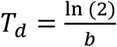. This is a statistical, but not an epidemiological model, that is, we are neither trying to model infection transmission nor estimate epidemiological parameters, such as the pathogen’s reproductive or attack rate. Instead, we are fitting a curve to observed outbreak data at the country, state, and county level. A change-point analysis using the Fisher discriminant ratio as a kernel function does not show any significant change points in the outbreak, and therefore justifies modeling the COVID-19 outbreak as a phenomenon of unrestricted population growth.^8^ Because outbreak rates change over time and their estimation is somewhat sensitive to the starting figure,^4^ we alternatively calculate the outbreak rate after it reached 10 and 25 cases in the respective unit, finding a high correlation among the rates. We are aware that testing differences between states may also be important confounds. As the number of tests administered and the number of confirmed cases correlates to varying extents,^9^ this is however difficult to control for. A disadvantage of this statistical approach is that we cannot forecast outbreak dynamics, though we do not require extrapolated data in our work.

## Statistical results

For the entire U.S., the outbreak rate is 0.172, which translates to a doubling time of *T*_*d*_ = 4.025 days (as of April 13, 2020). At the state level, the average outbreak rate is 0.197 (*T*_*d*_ = 3.518) and the median is 0.194. The outbreak rate ranges from 0.085 to 0.282 with a standard deviation of 0.039. The spaghetti lines in Figure 1 trace the cases as a percentage of the maximum number of cases reported on April 13 at both levels. Across states, the outbreak rates follow a normal distribution, as evidenced by a Shapiro-Wilk test with *W* (51) = 0.991, *p* = 0.970 (Figure 3). We only identify Nebraska (outbreak rate = 0.085) as a potential outlier. To appropriately report the outbreak at the county level, we first remove all 869 counties where the outbreak has not yet commenced, that is where the growth rate is close to zero or where the number of reported cases is below five. For the remaining counties, the average outbreak rate is 0.134, which translates to *T*_*d*_ = 5.172 days. The median outbreak rate is 0.135, standard deviation 0.057, and the maximum 0.426 (Colonial Heights City, Virginia). Figure 2 shows the spaghetti lines for the state of South Carolina and its counties. The outbreak significantly deviates from a normal distribution, *W* (3145) = 0.982, *p* < 0.001 (Figure 4).

**Figure 1:**
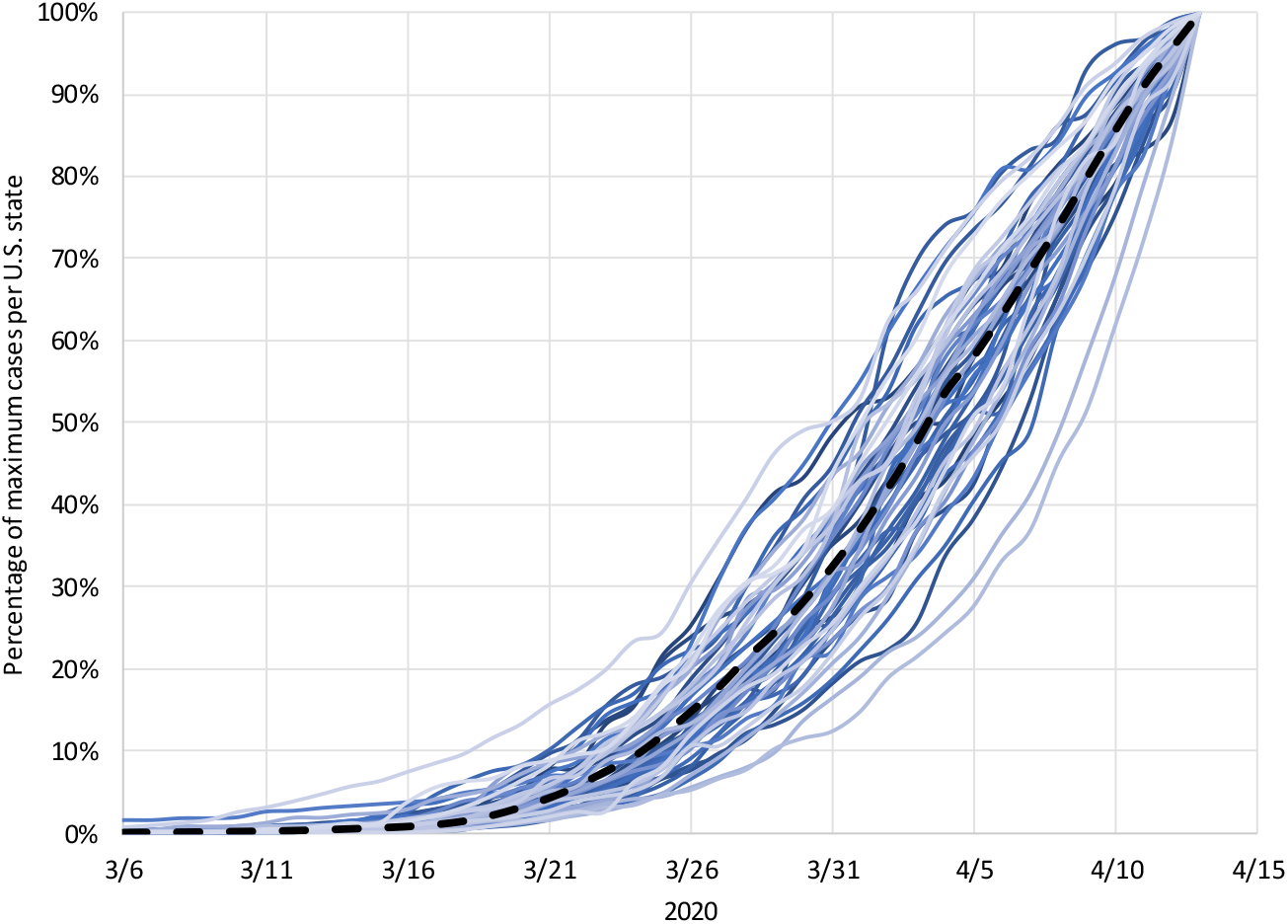
Epidemic days at U.S. country and state level. The spaghetti lines trace the COVID-19 outbreak in the U.S. (black dashed line) and the states (straight lines) as a percentage of the cases reported on April 13, 2020 in the entire country and each state, respectively.

**Figure 2:**
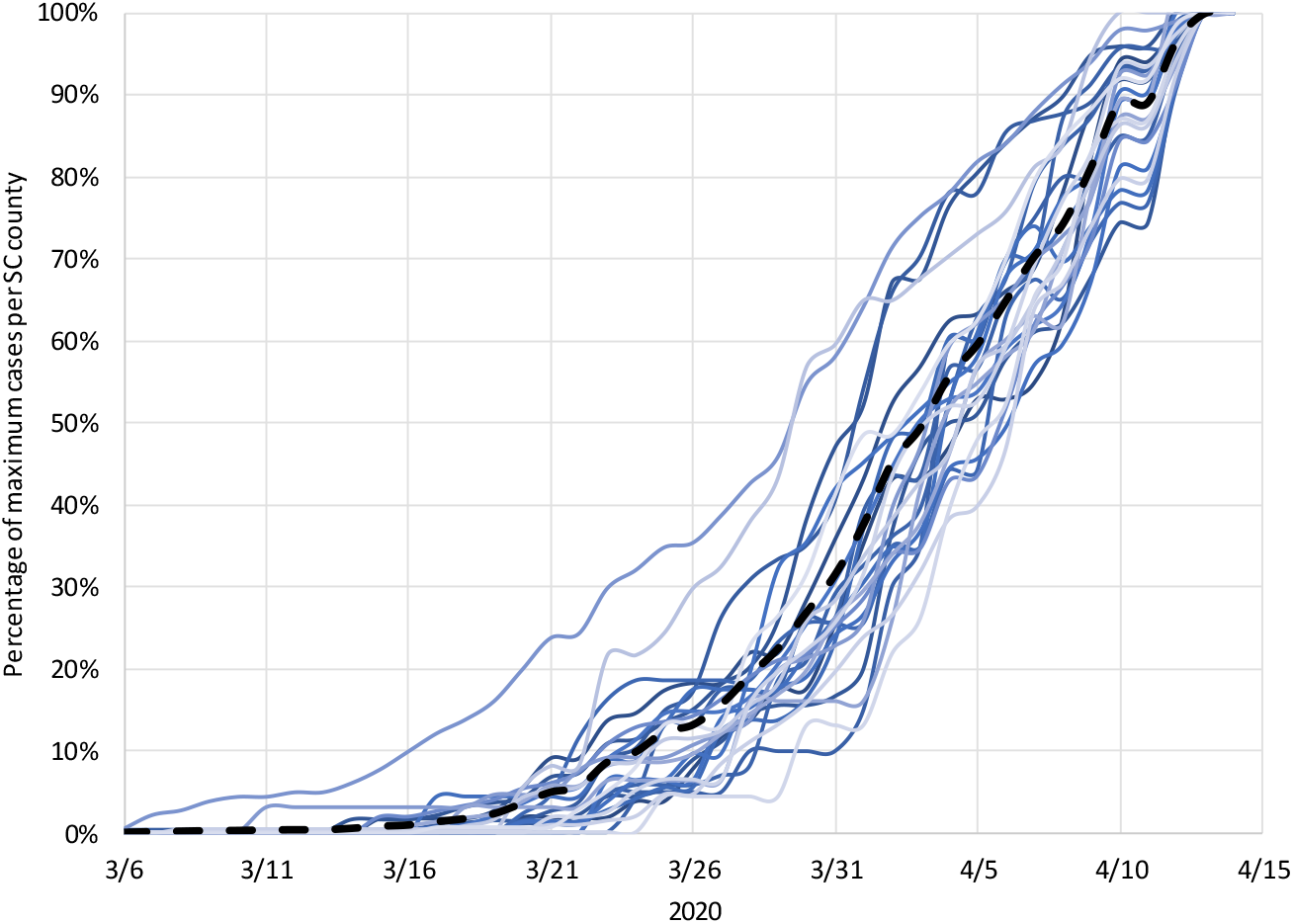
Epidemic days at county level (South Carolina) The spaghetti lines trace the COVID-19 outbreak in South Carolina (black dashed line) and the counties (straight lines) as a percentage of the cases reported on April 13, 2020. Cases unallocated to a county due to lack of information are included in the state line; counties with less than 20 reported cases on April 13 are not shown in the diagram.

**Figure 3:**
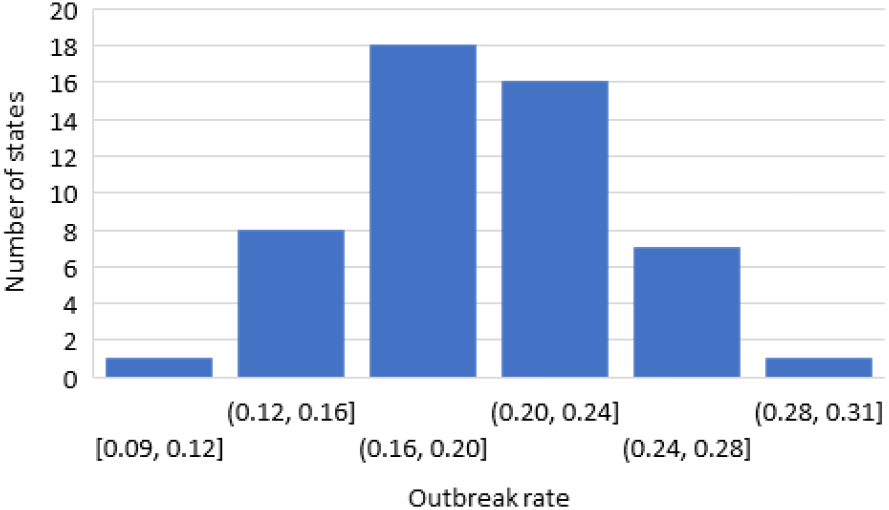
Distribution of outbreak rates across U.S. states. This histogram shows a normal distribution of the COVID-19 outbreak rate across 50 states plus Washington, D.C.

**Figure 4:**
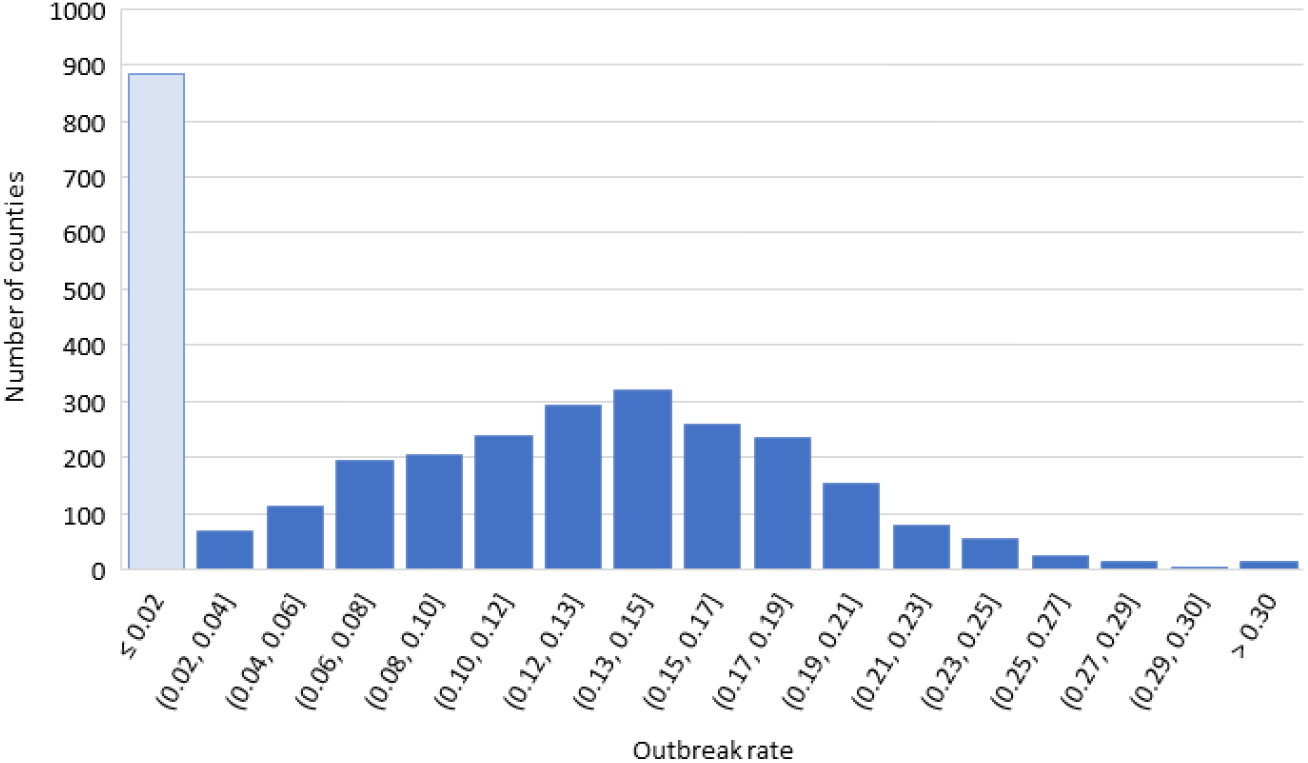
Distribution of outbreak rates across U.S. counties. This histogram shows the distribution of the COVID-19 outbreak rate across 3145 counties. The bar on the left captures all counties without an outbreak or a minimal number of cases.

The two geo maps in Figure 5 and Figure 6 show how the outbreak varies between states and between counties within a state. Statistically, we also find a very low degree of reliability between the state-wide and county-specific breakout rates. Even after removing counties without occurrence of the outbreak, the average measure ICC is very low at 0.169 with a 95% confidence interval from -0.080 to 0.360, *F*(2281,2281) = 1.462, *p* < 0.001. Similarly, Spearman’s ρ = 0.234 and Kendall’s τ = 0.160, both *p* < 0.001.^10^ The Bland-Altman plot shows a bias of 0.070 (standard error 0.001; standard deviation 0.062).^11^ In the same way, the two geo maps in Figure 7 and Figure 8 display the variation in time taken until the first ten cases were recorded on a state and county level. There is a larger variation in time-to-threshold at the U.S. county level as compared to the state level. Nebraska was the first to reach this threshold on February 17, 2020, with Texas, California, and Washington thereafter. In contrast, at the county level, King County (Washington) reached this threshold on March 02, Santa Clara (California) on March 03, Westchester County (New York) on March 04, and Los Angeles County (California) and Snohomish County (Washington) both on March 05. Yet, a significant portion of counties do not (yet) have a significant outbreak as of April 13 (Figure 4).

**Figure 5:**
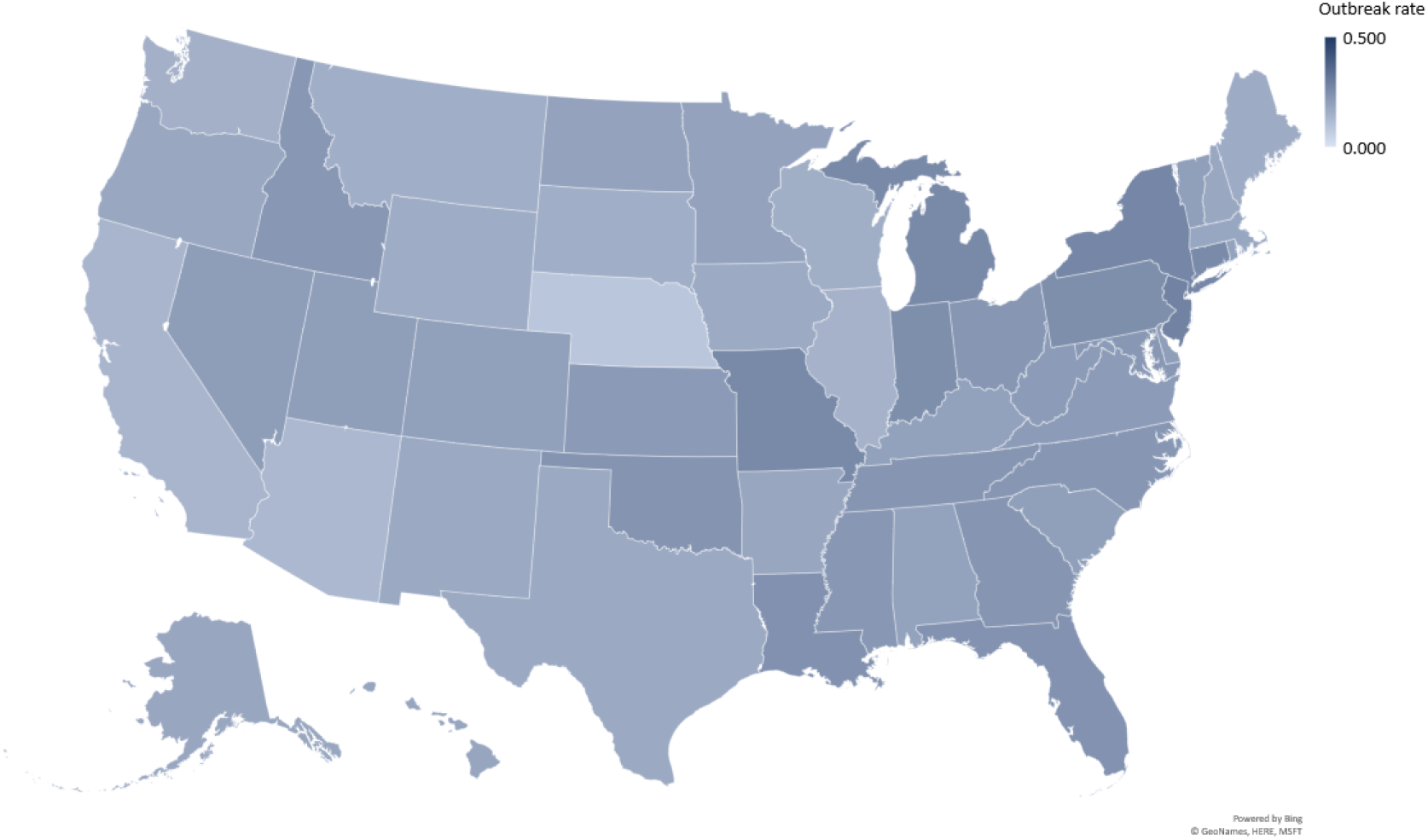
Variation in outbreak rates at U.S. state level. This geo map displays the variation in outbreak rates at U.S. state level. Lighter colors signify that the pandemic has a slower relative growth rate, and darker colors point to a faster growth.

**Figure 6:**
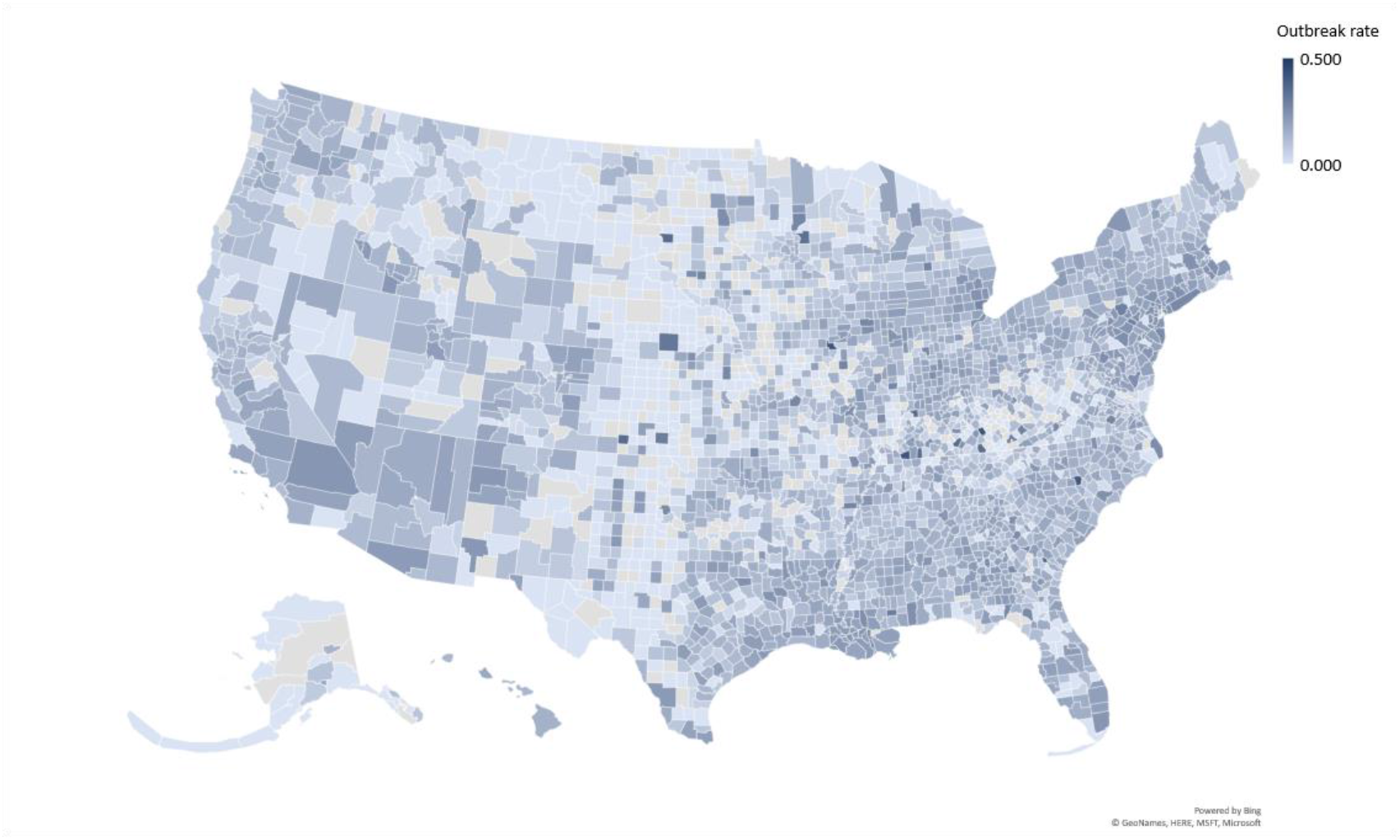
Variation in outbreak rates at U.S. county level. This geo map reveals the larger variation in outbreak rates at U.S. county level. The color band is the same as in Figure 5.

**Figure 7:**
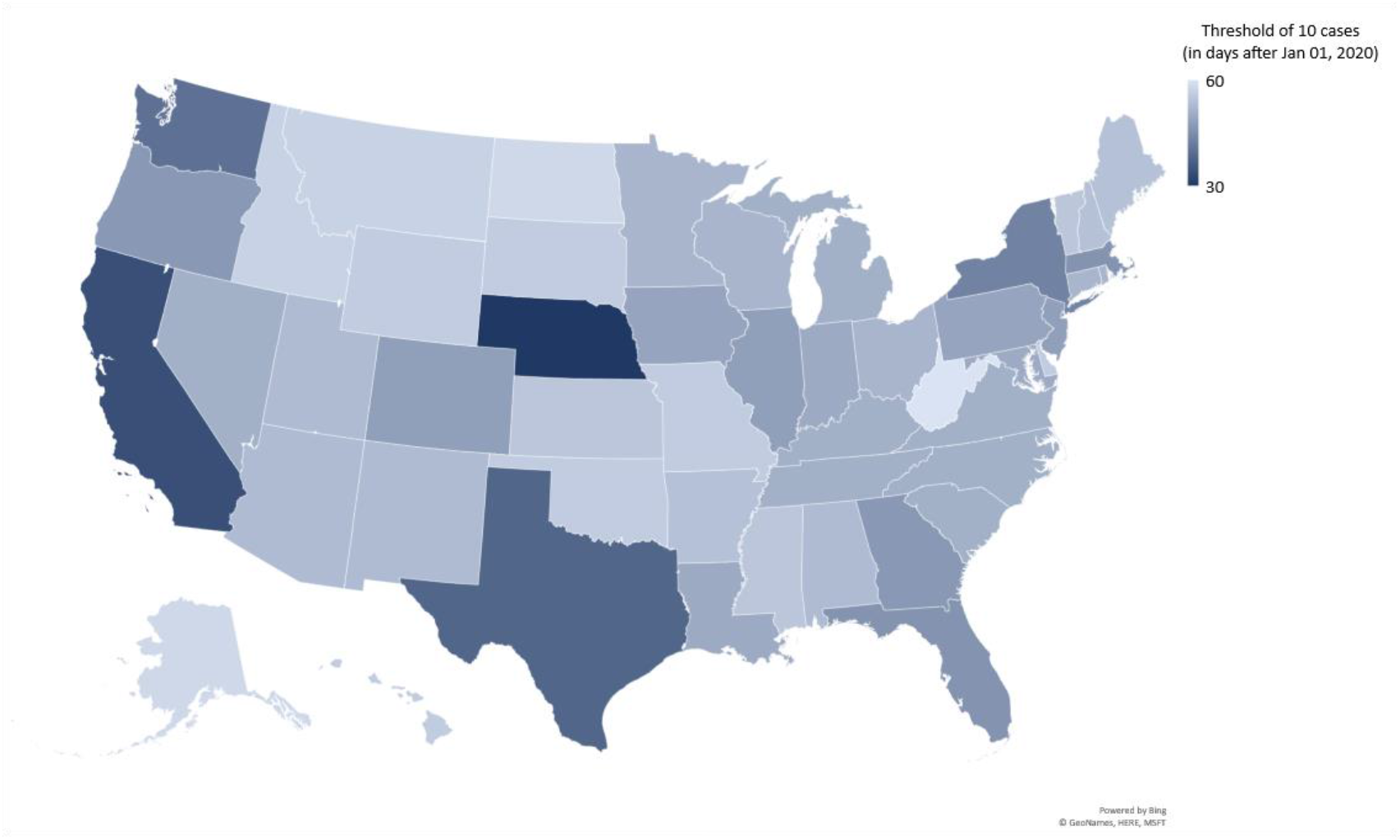
Variation of time to ten-cases threshold at U.S. state level. This geo map displays the variation in time taken till the first ten cases were recorded. Darker colors show states that have reached this threshold faster.

**Figure 8:**
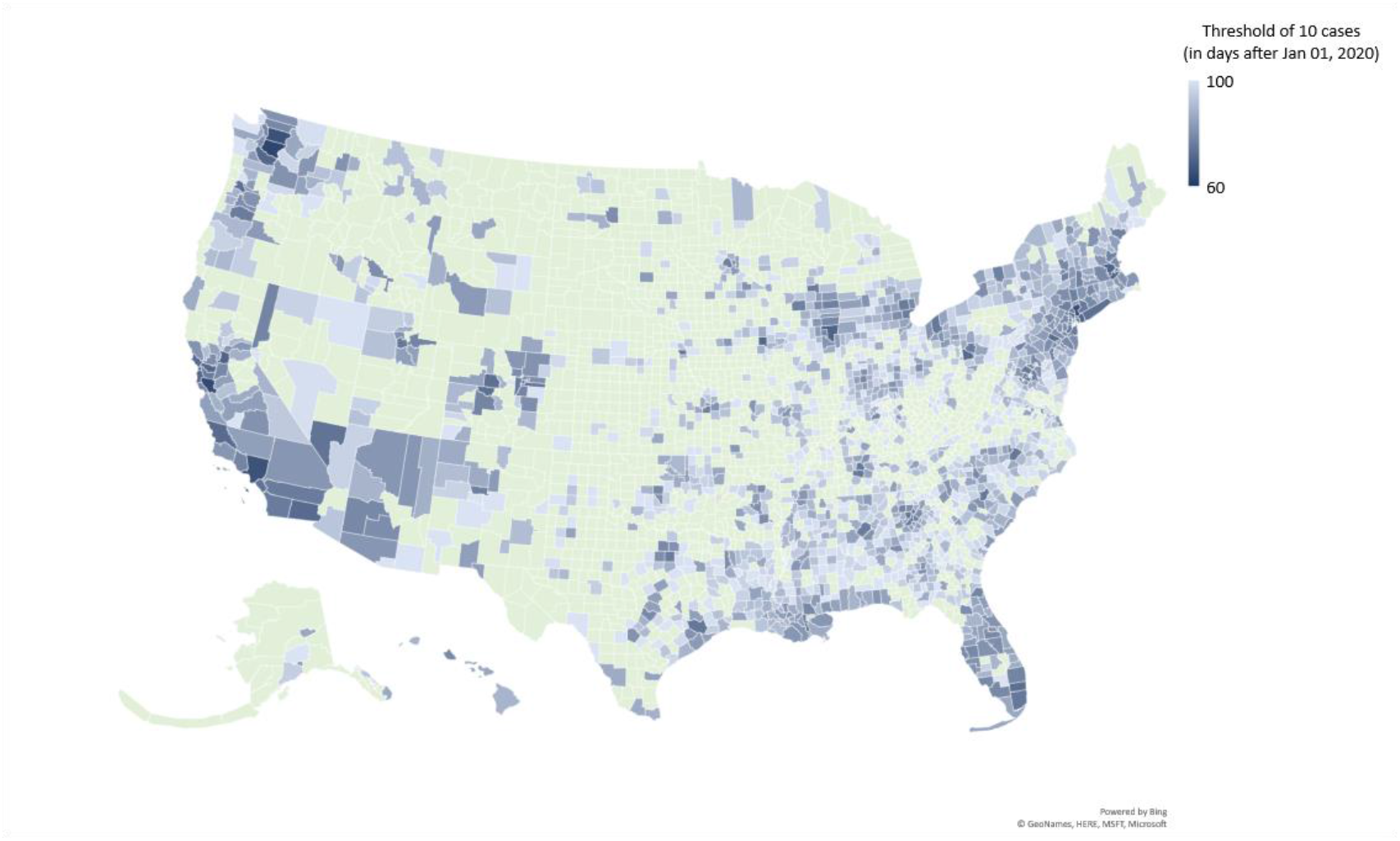
Variation of time to ten-cases threshold at U.S. county level. This geo map shows a larger variation in time-to-threshold at the U.S. county level as compared to the state level (Figure 7). Darker blue colors show counties that have reached this threshold faster. Counties without a significant outbreak as of April 13, 2020 are coded in light green.

## Discussion

In the U.S., the outbreak of the COVID-19 pandemic varies considerably not only between states, but also within the counties of a state. The histogram in Figure 3 expresses a normal distribution of the COVID-19 outbreak rate across 50 states plus Washington, D.C. When we extrapolate this to the county data, we find that the outbreak data significantly deviates from a normal distribution, even when omitting the counties with little to no outbreak (Figure 4). When graphed, this variation in case counts from county to county is easily visible (Figure 2 and Figure 6). In comparison with state level depiction (Figure 5), there is great variation between the state ranking and the situation in its individual counties. In the U.S., most response measures to the pandemic are devised and effected at the state level. Although this is certainly better targeted than an overall response at the federal level, which might spread resources too thinly in some regions, it still may not cater sufficiently for local outbreak differences and resource utilization. For example, while many counties in South Carolina still conveyed a utilized hospital bed capacity of less than 50% (as of April 21, 2020), Lexington County reported 90.6%, followed by Orangeburg and Colleton Counties at 82.2 and 78.0% respectively.^12^

Political policies play a large role in the resolution of national health emergencies, and have been found to be the strongest predictor of the early adoption of social distancing policies.^13^ But such policies tend to generalize strategy and target larger populations. Various institutional, societal, and cultural factors influence the development and adoption of these policies, and are important in the analysis of variations in the pandemic’s growth rate across states and counties. Between countries at the international level, previous research indicates the association of such contextual factors with the outbreak rate.^14^ For the U.S., we expect comparable findings, and aim to understand potential reasons for the differences in further research.

More generally, our study indicates that governments must track a pandemic’s outbreak and tailor appropriate response strategies to the most granular level possible. This would not only increase effectiveness of political policy and response strategy, but also allow for a redistribution of excess resources to areas most vulnerable to the pandemic. This will become increasingly important as the world begins returning to normalcy, and attempts to prevent further waves of the COVID-19 pandemic.

## Human participant protection

No humans participated in this study. The original data sources are referenced in the section Model and methods.

## Data Availability

The original data sources are referenced in the section Model and methods.

## Funding and acknowledgement

We gratefully acknowledge support through the Darla Moore School of Business and the Center for International Business Education and Research (CIBER) at the University of South Carolina.

## References

1. China Data Lab. US COVID-19 daily cases with basemap. Harvard Dataverse. https://doi.org/10.7910/DVN/HIDLTK. Published 2020. accessed April 14, 2020.

2. Chen JT, Kahn R, Li R, et al. U.S. county-level characteristics to inform equitable COVID-19 response. medRxiv. 2020. doi:10.1101/2020.04.08.20058248

3. Lang A. Why is the coronavirus death rate higher in Horry County than most of South Carolina? Myrtle Beach Online. https://www.myrtlebeachonline.com/news/coronavirus/article241915311.html. Published 2020. accessed April 22, 2020.

4. White ER, Hébert-Dufresne L. State-level variation of initial COVID-19 dynamics in the United States: The role of local government interventions. medRxiv. 2020:1–14. doi:10.1101/2020.04.14.20065318

5. USA Facts. Coronavirus locations: COVID-19 map by county and state. https://usafacts.org/visualizations/coronavirus-covid-19-spread-map/. Published 2020. accessed April 14, 2020.

6. Murray CJ. Forecasting COVID-19 impact on hospital bed-days, ICU-days, ventilator-days and deaths by US state in the next 4 months. medRxiv. 2020:1–26. doi:10.1101/2020.03.27.20043752

7. Woody S, Tec M, Dahan M, et al. Projections for first-wave COVID-19 deaths across the U.S. using social-distancing measures derived from mobile phones measures derived from mobile phones. The University of Texas at Austin COVID-19 Modeling Consortium. https://www.tacc.utexas.edu/ut_covid-19_mortality_forecasting_model_report. Published 2020. accessed April 19, 2020.

8. Texier G, Farouh M, Pellegrin L, et al. Outbreak definition by change point analysis: A tool for public health decision? BMC Med Inform Decis Mak. 2016;16(1):1-12. doi:10.1186/s12911-016-0271-x

9. Kaashoek J, Santillana M. COVID-19 positive cases, evidence on the time evolution of the epidemic or an indicator of local testing capabilities? A case study in the United States. SSRN. 2020:1–14. doi:10.2139/ssrn.3574849

10. Marra CA, Lynd LD, Harvard SS, Grubisic M. Agreement between aggregate and individual-level measures of income and education: A comparison across three patient groups. BMC Health Serv Res. 2011;11:1–7. doi:10.1186/1472-6963-11-69

11. Bland MJ, Altman DG. Measuring agreement in method comparison studies. Stat Methods Med Res. 1999;8(2):135-160. doi:10.1177/096228029900800204

12. SCDHEC. Hospital bed capacity (COVID-19). https://www.scdhec.gov/infectious-diseases/viruses/coronavirus-disease-2019-covid-19/hospital-bed-capacity-covid-19. Published 2020. accessed April 22, 2020.

13. Adolph C, Amano K, Bang-Jensen B, Fullman N, Wilkerson J. Pandemic politics: Timing state-level social distancing responses to COVID-19. medRxiv. 2020:1–19. doi:10.1101/2020.03.30.20046326

14. Messner W. The institutional and cultural context of cross-national variation in COVID-19 outbreaks. medRxiv. 2020:1–13. doi:10.1101/2020.03.30.20047589

